# Childhood coordination and survival up to six decades later: extended follow-up of participants in the National Child Development Study

**DOI:** 10.1101/19004713

**Authors:** G. David Batty, Ian J. Deary, Mark Hamer, Stuart J. Ritchie, David Bann

**Affiliations:** Department of Epidemiology & Public Health, University College London, UK; School of Biological & Population Health Sciences, Oregon State University, USA; Centre for Cognitive Ageing & Cognitive Epidemiology, University of Edinburgh, UK; School of Sport, Exercise & Health Sciences, Loughborough University, UK; Social, Genetic & Developmental Psychiatry Centre, King’s College London, UK; Centre for Longitudinal Studies, University College London Institute of Education, UK

**Keywords:** cohort, coordination, developmental origins of adult disease, epidemiology, mortality

## Abstract

**Background:** Poorer performance on standard tests of motor coordination in children has emerging links with sedentary behaviour, obesity, and functional capacity in later life. These observations are suggestive of an as-yet untested association of coordination with health outcomes.

**Objective:** To examine the association of performance on a series of psychomotor coordination tests in childhood with mortality up to six decades later.

**Design, Setting, and Participants:** The National Child Development Study (1958 birth cohort study) is a prospective cohort study based on a nationally representative sample of births from England, Scotland and Wales. A total of 17,415 individuals had their gross and fine motor psychomotor coordination assessed using nine tests at 11 and 16 years of age.

**Main outcome and measure:** All-cause mortality as ascertained from a vital status registry and survey records.

**Results:** Mortality surveillance between 7 and 58 years of age in an analytical sample of 17,336 men and women yielded 1,090 deaths. After adjustment for sex, higher scores on seven of the nine childhood coordination tests were associated with a lower risk of mortality in a stepwise manner. After further statistical control for early life socioeconomic, health, cognitive, and developmental factors, relations at conventional levels of statistical significance remains for three tests: ball catching at age 11 (hazard ratio; 95% confidence interval for 0-8 versus 10 catches: 1.56; 1.21, 2.01), match-picking at age 11 (>50 seconds versus 0-36: 1.33; 1.03, 1.70), and hopping at age 16 years (very unsteady versus very steady: 1.29; 1.02, 1.64).

**Conclusion and Relevance:** The apparent predictive utility of early life psychomotor coordination requires replication.

**Key points:** *Question:* What is the association of performance on a series of psychomotor coordination tests in childhood with mortality up to six decades later?

*Findings:* After taking into account multiple confounding factors, lower performance on three gross and fine motors skills tests in childhood were associated with a shorter survival over six decades.

*Meaning:* These findings require replication in other contexts and using complementary observational approaches.

## Introduction

Coordinated movement is the product of the dynamic and complex interplay of multiple neural mechanisms that are modulated by sensory inputs and spinal reflex loops.^1^ While there is a literature on the social and educational sequela of poorer performance on standard tests which capture the capacity to coordinate movement,^2,3^ there is growing evidence for a detrimental impact on health-related outcomes in later life. In extended follow-up of general population based surveys into middle- and older age, poorer motor skill scores in childhood that nonetheless lie within the normal range are associated with future physical inactivity,^4,5^ lower functional capacity,^6^ higher body mass index,^7^ mental health problems,^8,9^ and worse self-rated health.^9^ Thus, findings from a national birth cohort study, for instance, suggest that higher scores on fine motor coordination tasks at age 15 years were related to better performance on physical function tests of standing balance and chair rising at age 53 years^6^ – test results that themselves have been linked to greater life-expectancy.^10^ In a similar longitudinal study, lower scores on tests at age 11 years were associated with a higher risk of obesity two decades later.^7^ Linear effects were evident across the full range of coordination scores such that the association was not generated by children at the lowest end of the continuum in whom a diagnosis of coordination disorder was likely. In comparisons of the characteristics of children with such a diagnosis, relative to unaffected controls, there are also reports of a greater likelihood of raised levels of blood pressure and triglycerides, less favourable cardiac output, and lower left ventricular volume.^11,12^

Taken together, these findings provide a *prima facie* case for the hypothesis that poorer scores on tests of psychomotor coordination in early life may be associated with premature mortality in adulthood. We tested this proposition using a six decade follow-up of over seventeen thousand members of the 1958 British birth cohort study. Given that children with lower coordination scores may come from disadvantaged social backgrounds and have less favourable levels of cognitive function and birth characteristics—all of which are pre-adult risk factors for mortality in their own right^13,14^—we took these factors into account, thus exploring whether there was an independent association of mortality with earlier psychomotor coordination.

## Methods

Described in detail elsewhere,^15^ the National Child Development Study (or the 1958 birth cohort study) is an ongoing, prospective birth cohort study initially comprising 17,415 births to mothers residing across Britain (England, Wales, Scotland) in one week of 1958. Following data collection during the perinatal period, to date, there have been ten follow-up surveys of study members in the National Child Development Study.^15^ These have been designed to monitor their physical, educational, social, and health development up to age 55 years. Responders remain broadly representative of the original sample.^16^ Ethical approval has been granted by the South-East Multi-Centre Research Ethics Committee, and study participants provided informed consent.

### Assessment of early life physical coordination

At 7 years of age, qualitative judgments about the study member’s motor skills were sought from the teacher or parent, whereas at 11 years and 16 years, psychomotor tests were administered by a school medical officer.^17^ Owing to the subjective nature of the enquiries at age 7, our main results are for the later tests which included fine and gross motor skill evaluations (for the age 7 results, see the supplemental figure 1).

At age 11 years, we utilised results from 6 tests of motor coordination. Children were asked to stand on their right foot for 15 seconds, to stand heel-to-toe for 15 seconds, and to walk backwards along a line, hands on hips, toe-to-heel, for 20 steps. Performance on these tests was categorised into one of three groups (‘very steady’, ‘slightly unsteady’, or ‘very unsteady’). Next, standing with their forearm horizontal, study members were instructed to bounce a ball, catching it with palm facing downward; the number of catches out of 10 attempts was recorded (categories: 0-8, 9, 10 catches [best]). Participants were also asked to individually pick up and transfer 20 matches from one matchbox to another; the time taken was recorded and categorised into quartiles for the purposes of the present analyses (0-36 seconds [fastest], 37-43, 44-50, >50). Lastly, on a printed grid of 200 squares, the children were asked to mark as many squares as possible within 1 minute; again, these data were categorised into quartiles: (>126 squares [highest], 110-126, 95-109, 0-94). From age 16 years, we utilised three coordination tests. The heel-toe standing balance and ball catching test were re-administered together with a hopping test. In the latter test, participants jumped one legged between four lines spaced two feet apart, swivelled, then returned on the same leg. Performance was rated as very steady, slightly unsteady, and very unsteady.

These coordination tests, amongst the earliest to be developed, were subsequently incorporated into batteries in common usage today, including the Zurich Neuromotor Assessment,^18^ the Movement Assessment Battery for Children,^19^ and the McCarron Assessment of Neuromuscular Development.^20^ Inter-rating reliability and test-retest reliability of these test batteries are high, scores correlate moderately well with reaction time, and have diagnostic utility.^21^

### Assessment of potential confounding factors

Birth weight was recorded by the midwives. Gestational age was based on the number of days from the first day of the last menstrual period. At birth, enquiries were made about the occupational social class of the mother and father (six categories from professional to unskilled),^22^ maternal attendance in post-compulsory school, overcrowding, maternal weight (1 stone is 14 pounds), and smoking status. When the study member was 7 years of age, mothers also responded to questions about whether the child had been breastfed, and if they thought their child was right-handed, left-handed, or ambidextrous (mixed handed). Height and weight were measured by trained medical personnel at age 7 using standardized protocols,^23^ and study members were administered the Southgate Group Reading Test and Problem Arithmetic Test.^24^

### Mortality ascertainment

Vital status was ascertained up to 58 years of age (December 2016) using death certificates supplied by the Office for National Statistics and/or notifications given during fieldwork following direct contact with participants’ family members.^25^ All-cause mortality from the date of coordination test administration (7, 11, or 16 years) to age 58 years was used in analyses.

### Statistical analyses

We summarised the relation between each test of coordination and mortality using Cox proportional hazards regression model.^26^ The proportional hazard assumption was tested by both visually inspecting hazard estimates and by testing Schoenfeld Residuals;^27^ there was no strong evidence for violation. Hazard ratios with accompanying 95% confidence intervals were computed and initially adjusted for gender (no age adjustment was required given that study members are aged within 7 days of one another), followed by potential confounding variables. To reduce the impact of missing data on statistical power and the occurrence of selection bias,^28^ missing data were generated using multiple imputation with 10 such datasets.

## Results

We first ascertained if psychomotor coordination test scores at 11 and 16 years were linked to study covariates. Taking results from one of the tests as an illustration, ball catching test results at age 11 were associated with cognitive ability, laterality, and health (table 1), such that children with lower coordination scores performed less favourably in these domains; however, for most of the potential covariates there was no evidence of a relationship. A somewhat dissimilar pattern of association was evident at age 16 years where, additionally, a higher prevalence of socioeconomic disadvantage (parental manual social class, overcrowding), an absence of breast feeding, and increased maternal and study members’ own weight was apparent in children who were less successful on the hopping test (supplemental table 1).

**Table 1.** Study member characteristics according to ball catches at age 11 years.

| Characteristic (age measured) | Number of ball catches<br>(N, %) |  |  |  | P-value for<br>heterogeneity |
| --- | --- | --- | --- | --- | --- |
|  | N | 10 (best) | 9 | 8-0 |  |
| Male | 12583 | 5068 (49.7) | 845 (56.1) | 533 (60.4) | <0.001 |
| Manual parental occupational social class (0 years) | 12547 | 7235 (71.2) | 1090 (72.6) | 650 (73.8) | 0.4 |
| Mother did not attend post compulsory school (0 years) | 11948 | 7153 (74.0) | 1044 (72.7) | 650 (77.3) | 0.2 |
| Overcrowding (>2persons/room; 0 years) | 11672 | 2896 (30.7) | 428 (30.5) | 251 (30.5) | 0.9 |
| Mother smoked prior to pregnancy (0 years) | 11640 | 3676 (39.0) | 518 (37.1) | 303 (37.1) | 0.4 |
| Mother did not breastfeed (7 years) | 11257 | 2787 (30.5) | 415 (31.3) | 270 (34.6) | 0.1 |
| Mother's weight (0 years) >12st | 11727 | 1171 (12.3) | 161 (11.4) | 86 (10.5) | 0.7 |
| Participants' weight >60lbs (7 years) | 10778 | 1179 (13.5) | 158 (12.4) | 96 (12.7) | 0.5 |
| Participants' height <45 inches (7 years) | 10966 | 479 (5.4) | 90 (7.0) | 71 (9.4) | <0.001 |
| Birth weight <2.5kg (0 years) | 11601 | 519 (5.5) | 81 (5.8) | 59 (7.2) | 0.1 |
| Gestational age <9 months (0 years) | 10814 | 1899 (21.6) | 254 (20.0) | 160 (21.1) | 0.4 |
| Cognition, low reading test (7 years) | 11423 | 1008 (10.9) | 199 (14.6) | 165 (20.8) | <0.001 |
| Cognition, low maths test (7 years) | 11389 | 2409 (26.1) | 400 (29.6) | 306 (38.4) | <0.001 |
| Pubertal timing, advanced (7 years) | 8170 | 2258 (34.2) | 349 (35.6) | 206 (34.8) | 0.9 |
| Laterality – left or mixed handed (7 years) | 11336 | 820 (8.9) | 195 (14.6) | 147 (18.6) | <0.001 |
| Health condition, chronic developmental (7 years) | 10962 | 874 (9.8) | 138 (10.7) | 116 (15.3) | <0.001 |
| Health condition, chronic disease/condition (7 years) | 10968 | 556 (6.2) | 71 (5.5) | 51 (6.8) | 0.5 |

P-values derived from chi-squared tests (overall heterogeneity); p-values calculated using all available categories, and binary proportions shown to aid presentation.

Mortality surveillance between 7 and 58 years of age in a maximum of 17,336 men and women yielded 1,090 deaths (835,714 person-years at risk). With the exception of mother’s weight, study member’s weight in childhood, gestational age, and laterality, the remaining thirteen covariates were related to mortality rates in the expected directions in univariate analyses (figure 1). Of the statistically significant relationships, the strongest association was evident for childhood cognition, the weakest for an absence of breast feeding.

**Figure 1.**
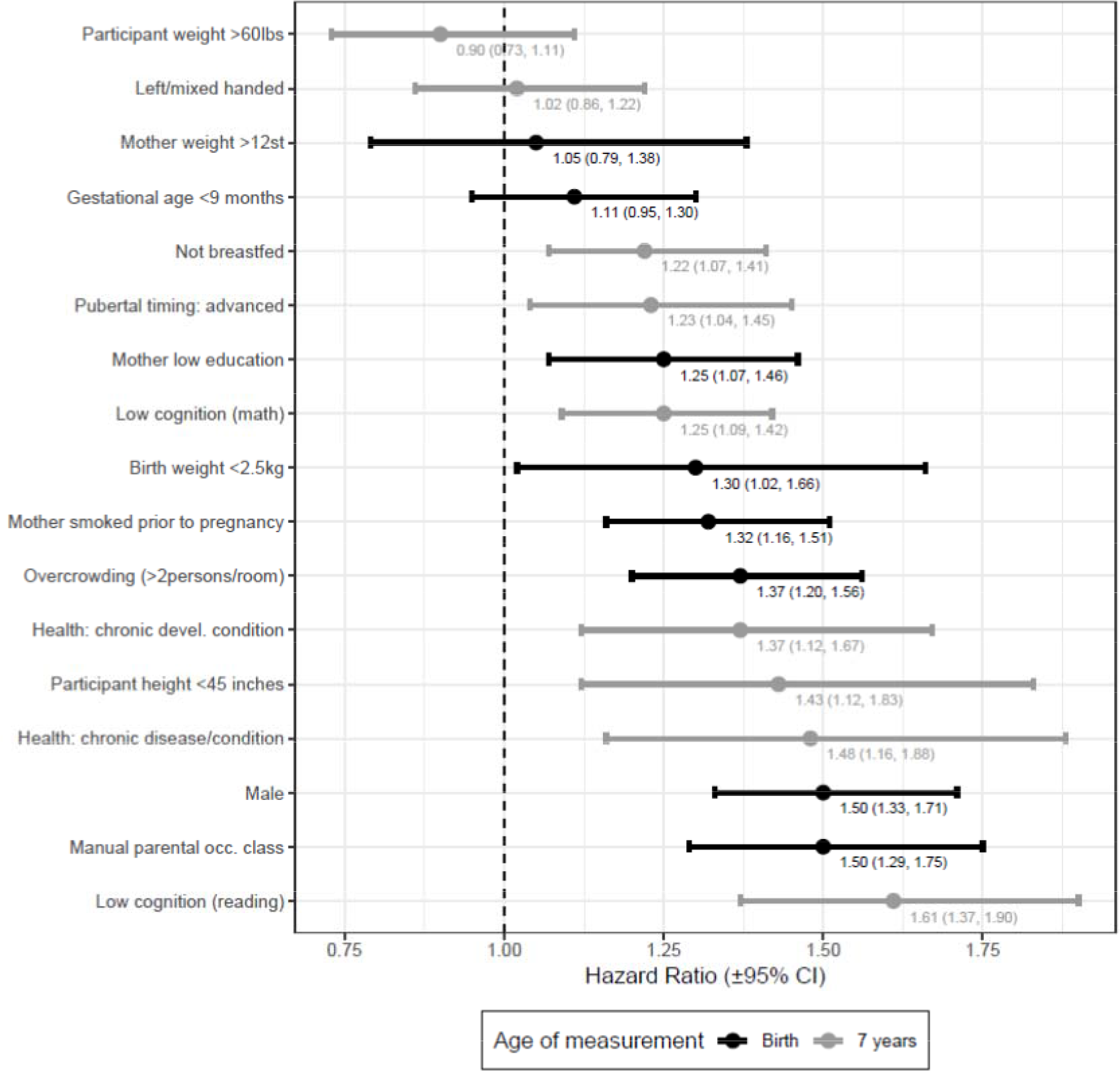
Early life characteristics in relation to total mortality up to age 58 years. Hazard ratios are unadjusted.

In figure 2 we show survival curves according to scores on the nine coordination tests administered in childhood (figure 2a) and adolescence (figure 2b). In these univariate analyses, lower survival rates were typically apparent in children and adolescents with poorer coordination test results. The clearest differences in survival across the full duration of mortality follow-up were apparent for ball-catching at 11 years of age, whereas heel-to-toe balance was not linked to earlier death. A composite indicator of coordination, constructed from a summation of the three numerically scored tests at age 11 (match-picking, square marking, ball catching) revealed similar results.

**Figure 2.**
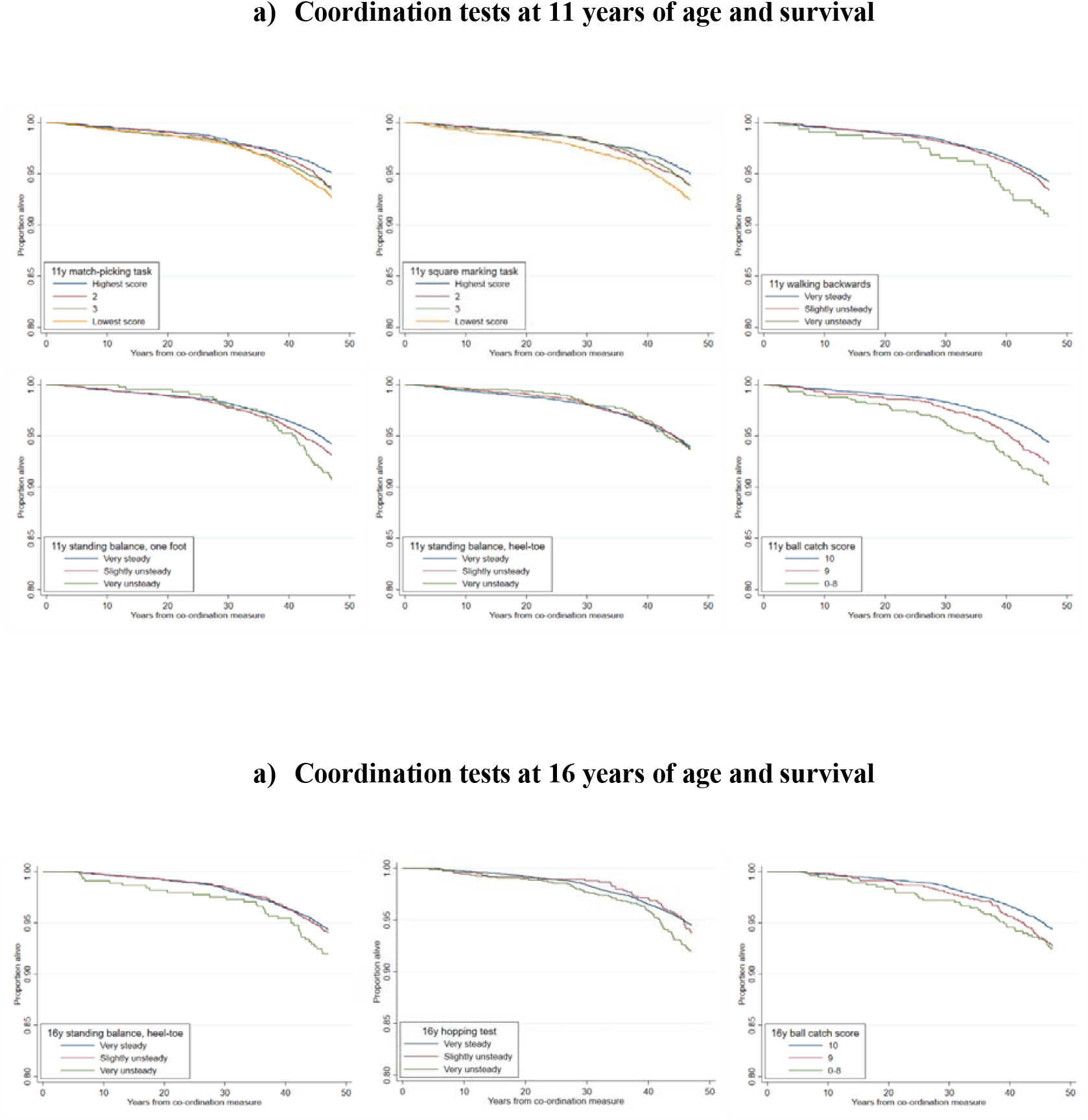
Kaplan–Meier survival curves according to results from coordination tests administered at (a) 11, and (b) 16 years of age.

Next, we expressed these findings as hazard ratios, while controlling for potentially confounding variables that might account for all or part of the coordination–death relation (figure 3 for tests administered at 11 years of age, figure 4 for tests at 16 years of age). In analyses with gender in the multivariable model, the majority of test scores from both time points—seven of the nine conducted—were inversely related to mortality risk, such that a higher concentration of deaths was apparent in children who had lower coordination scores. In the two tests that did not reveal a relationship with mortality risk – the standing balance heel-to-toe test at 11 and 16 years – the later test results were of borderline statistical significance. These effects were typically stepwise across the full range of coordination scores. The hazard ratios for the coordination tests at age 11 years were generally of higher magnitude than for those at age 16, although fewer tests were administered in adolescence. In analyses where 16 covariates were added to the multivariable model, there was clear attenuation across all relationships, and statistically significant associations were evident for only three tests (match-picking and ball catching at age 11 years, and hopping at 16 years).

**Figure 3.**
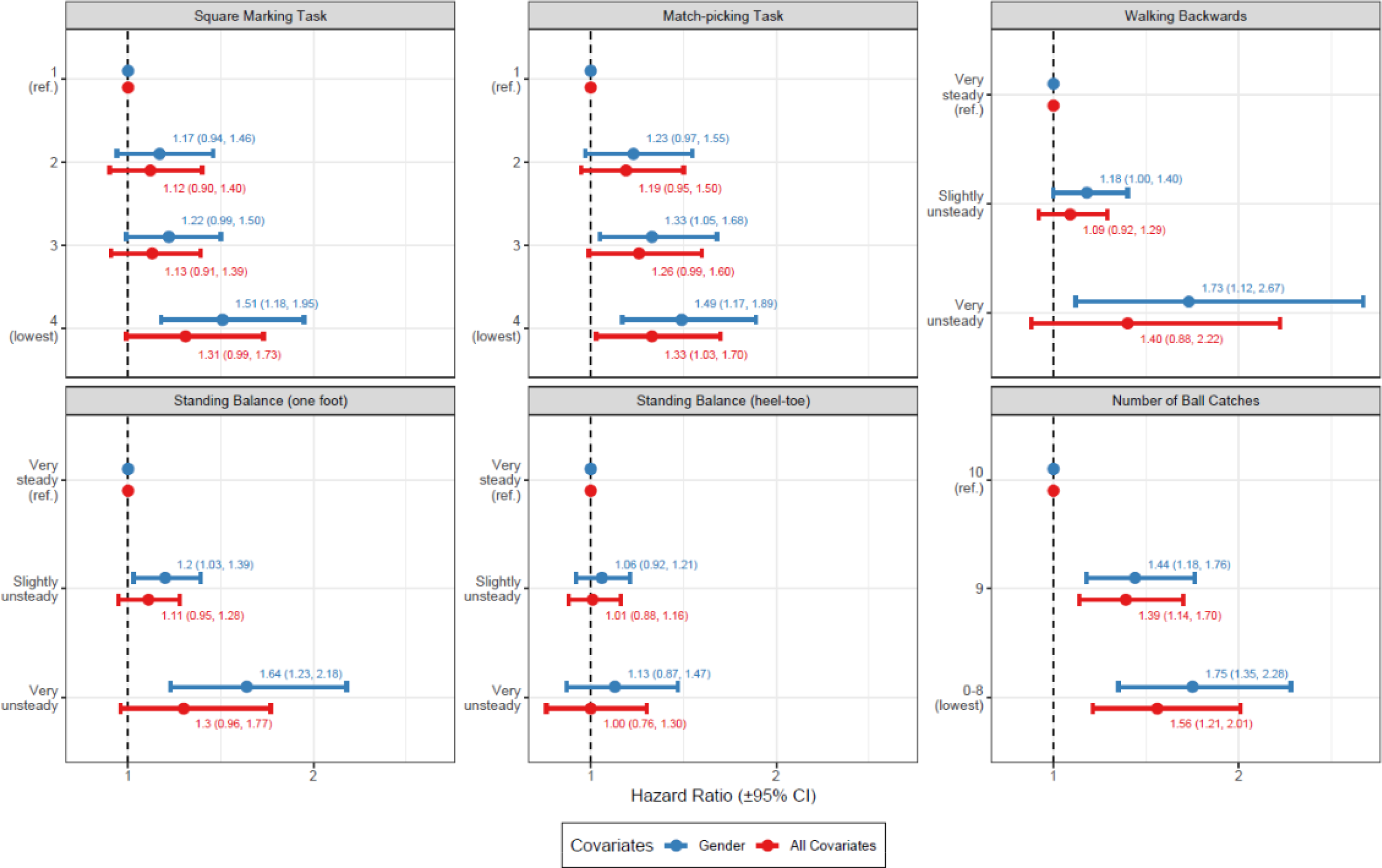
Multiply-adjusted hazard ratios (95 confidence intervals) for the relation of psychomotor coordination tests at age 11 years with all-cause mortality by 58 years. All Covariates comprise childhood socioeconomic, health, cognitive, and developmental factors as listed in table 1.

**Figure 4.**
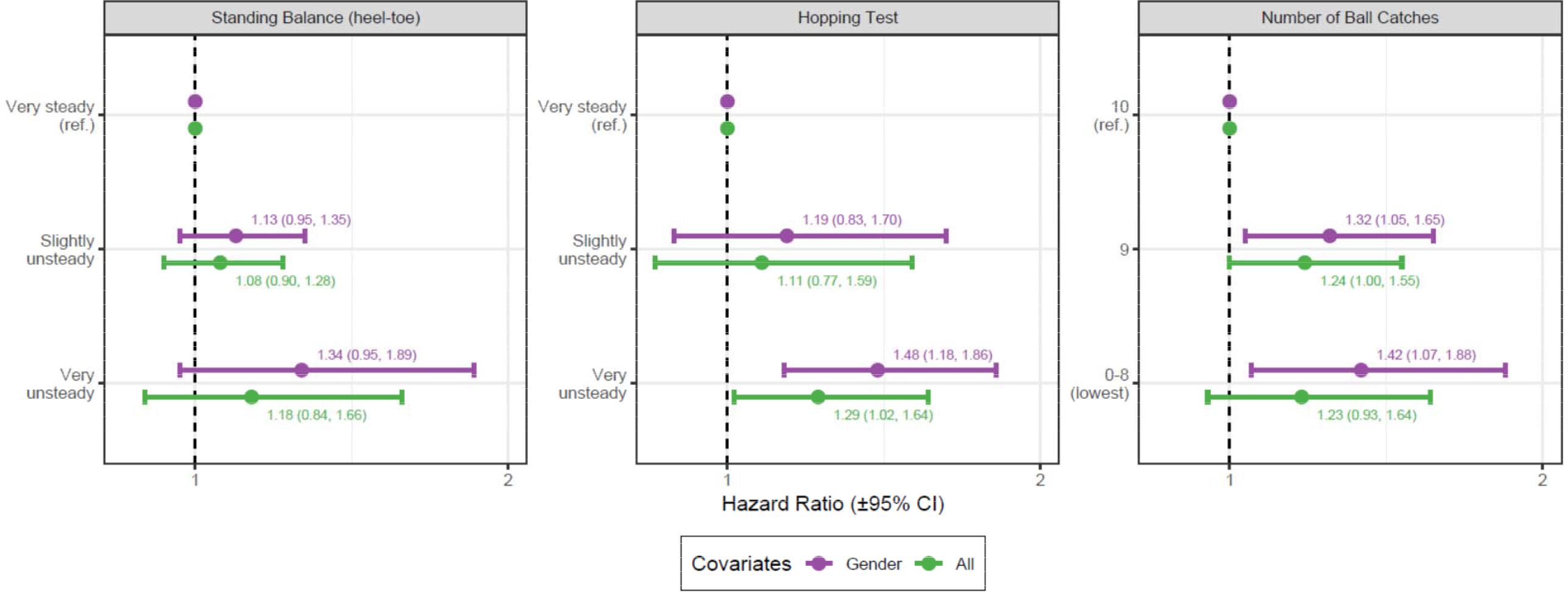
Multiply-adjusted hazard ratios (95 confidence intervals) for the relation of psychomotor coordination tests at 16 years with all-cause mortality by 58 years. All Covariates comprise childhood socioeconomic, health, cognitive, and developmental factors as listed in table 1.

The strongest relationship with mortality were evident with the parent or teacher’s subjective reporting of study member’s coordination at 7 years (supplemental figure 1) as opposed to the results from the tests administered at later ages. This maybe because the evaluations were more prone to the observer’s knowledge of existing psychomotor problems in the school children, including those with developmental coordination disorder.

## Discussion

The main findings of the present study was that better performance on the majority of childhood coordination tests results were associated with a lower risk of mortality up to six decades later. Most associations were graded across the test score categories. After further control for an array of socioeconomic, health, cognitive, and developmental factors, these relationships held at conventional levels of statistical significance for the ball catching, match-picking duration, and hopping tests. We were able to recapitulate established associations from other life course-orientated studies whereby the early life characteristics of socioeconomic position,^29,30^ cognitive function,^14,31,32^ birth characteristics,^13^ and post-natal growth^33,34^ were all related to the risk of death up to six decades later (figure 1). This therefore increases confidence in our novel results for psychomotor coordination.

Given the large number and array of covariates included in our statistical models, particularly in socioeconomic, cognitive, and health domains, it is likely that the reported hazard ratios are underestimates of the risk of lower levels of coordination. Despite this potential over-adjustment, there was still evidence of an elevated risk of death for poorer performance on three tests. We did not have data on physical activity in early life in the present study, but given the known association with both coordination and adult mortality,^35^ it is a candidate confounder. However, since there was still a mortality association with the match-picking test, which captures fine-motor skills, this effect is unlikely to be fully-ascribable to the impact of practice in sports and exercise.

### Comparison with other studies

To the best of our knowledge, this is the first study to test the hypothesis that early life tests of coordination are associated with survival into adulthood so direct comparison with extant results is not possible. Several of the tests of coordination conducted in childhood in the present study do, however, bear some useful resemblance to standard assessments of physical capacity used in populations of older adults.^36^ These include the one-leg and heel-to-toe balance tests and the walking test; poorer performance on these measures is related to earlier mortality and with similar magnitude to those in the present analyses.^10^ In these older populations, reduced performance on such tests is likely to be at least partially ascribed to a higher occurrence of co-morbidity (hypertension, stroke, cognitive impairment etc). This explanation is not likely to be relevant to a cohort of children, in whom such illnesses would generally be very rare at the time of the psychomotor assessment.

### Mechanisms of effect

The robustness to statistical adjustment of the coordination–death relationships for selected tests raise the possibility of direct and indirect mechanisms. Our outcome in these analyses was mortality from all-causes and, although we did not have data on individual causes of death, in higher income countries mortality by 60 years of age is known to broadly comprise chronic disease (chiefly cardiovascular disease and cancer) and external causes of death (chiefly suicide and accidents, particularly road traffic). We speculate that the association between childhood coordination and external cause of death such as road traffic accidents may be direct: superior coordination results may capture an ability to rapidly respond to threat by removing oneself from high-risk situations, whether in the position of pedestrian or vehicle driver, so minimising injury and preserving life.^37^ A further direct effect of coordination on mortality may be via ‘system integrity’ which may have more relevance to chronic disease. First posited in the context of intelligence,^17^ system integrity concerns the suggestion, as yet not widely explored, that scoring well on tests of psychomotor coordination might be an indicator of a more general tendency for complex systems to function optimally. Thus, children with higher test scores may also have superior functioning of vital organs such as heart, lung, liver, and kidneys and, as such, enhanced life expectancy.^38,39^

Other coordination–chronic disease effects may be indirect. That is, they might be at least partially mediated by characteristics known to be affected by coordination that are also established risk factors for disease. These might include lifestyle factors (e.g., diet)^40,41^ and their correlates such as obesity.^42,43^ Physical activity is unlikely to be one of these since both fine and gross motor skills were linked with mortality risk. Adult mental health problems, associated with earlier measurement of coordination,^9^ are related to mortality risk in their own right.^44-46^ Although we were interested in the influence, if any, of normal-range coordination scores on mortality risk, as described, there is a suggestion that, relative to unaffected controls, children with developmental coordination disorder, experience raised levels of blood pressure and triglycerides, less favourable cardiac output, and lower left ventricular volume.^11,12^ These mechanisms may also be implicated in children who scored sub-optimally on the coordination test, but who nonetheless did not meet the criteria for a diagnosis of developmental coordination disorder. Although some of these intermediary data were collected in middle-aged participants in the present study, the number of deaths in those with both childhood coordination *and* adult risk factor data is currently too few to facilitate reliable mediation analyses. Lastly, it is plausible that there is a genetic correlation between the known genetic variants linked to longevity^47^ and those that predispose participants to better coordination. Should a large-scale genome-wide association study of life expectancy be performed, this issue could be investigated using statistical genetic techniques.

### Conclusion

In the present study, fine and gross motor skills were both linked with mortality risk up to six decades later. These findings require replication.

## Data Availability

Data from the National Child Development Study are available for downloading from the UK Data Archive (https://data-archive.ac.uk/).

